# Reporting of pre-analytical Processes in Biobanked Biomaterials: A Missed Opportunity to Address the Replicability Crisis in Clinical Biomarker Research

**DOI:** 10.1101/2025.02.18.25322469

**Authors:** Jannes A.W. Jansen, Mariël A. van den Brand, Femmie de Vegt, Dorine W. Swinkels

## Abstract

**Objectives:** Biobanks are crucial for clinical biomarker research, but pre-analytical processing can impact sample suitability. Insufficient reporting of pre-analytical methods can lead to invalid conclusions.

**Methods:** This study evaluated the pre-analytical reporting quality in 294 clinical biomarker studies using biobank plasma and serum samples published from 2018 to 2023, based on the SPREC and BRISQ guidelines.

**Results:** The analysis revealed frequent incomplete reporting. Key underreported elements included fasting time (reported in 31% of articles), freeze-thaw cycles (22.8%), internal transport (8.5%), and centrifugation settings (20-35%). Demographic data (96.9%), storage temperatures (81%), and blood tube additives (82.7%) were more consistently reported. No significant correlation was found between reporting quality and journal impact factor.

**Conclusions:** Our findings highlight the need for standardized pre-analytical reporting to improve research replicability and biomarker study quality.

## Introduction

Replicability is a cornerstone of clinical research, as it ensures that research findings can be consistently reproduced when methodologies are transparently reported. However, Ioannidis^1^ observed that over 30% of highly published clinical research articles fail to replicate, yielding less promising or even contradictory results in subsequent studies. Efforts to reproduce previous preclinical research findings also reveal concerning trends; for instance, Amgen and Bayer independently reported reproducibility rates of just 25% and 11%, respectively.^2,3^ More recently, the Center for Open Science confirmed these findings, with replication success rates of only 46% for results from high-impact papers.^4^

Several factors contribute to this lack of reproducibility, with inadequate documentation and reporting being significant contributors.^5,6,7^ For example, Sun et al. ^8^ identified poor reporting of analytical characteristics of biomarkers used in clinical research. We propose that insufficient characterization and documentation of the pre-analytical phase for biobank biomaterials could be an underappreciated factor influencing research replicability and transparency.^9,10^

The pre-analytical phase encompasses all procedures conducted prior to the analysis of samples. It comprises factors of biological origin that may occur *in vivo* or *in vitro. In vivo* factors include effects of health and disease status and can be modifiable such as diet and circadian rhythm, or unmodifiable such as age, gender, race, and genetic background, *In vitro factors* include samples collection, processing, transport, and storage.

Accurate reporting of pre-analytical factors is essential to verify whether the stored materials accurately represent the biomaterials intended for analysis and to assess their fitness for the intended biomarker measurements. Numerous studies have demonstrated that errors in the pre-analytical phase, such as incorrect additives, delays between sample collection and centrifugation/storage, and suboptimal storage conditions, can significantly impact measurement outcomes.^9,10^ For instance, results from studies measuring (mi)RNA in plasma have been shown to be influenced by factors such as tube additives and storage temperature.^9,11^ Additionally, metabolomic profiles in plasma and serum are highly susceptible to pre-analytical variables, including fasting status, collection time, hemolysis, delays in processing, and repeated freeze-thaw cycles.^12-15^ Therefore, comprehensive reporting of sample collection and handling procedures is critical to ensuring the reliability and replicability of research findings.

Several guidelines have been developed to assist researchers in reporting pre-analytical factors. Notably, the Sample PREanalytical Code (SPREC) and the Biospecimen Reporting for Improved Study Quality (BRISQ) criteria have been established to standardize the reporting of pre-analytical procedures.^16-18^ SPREC focuses on metadata related to *in vitro* pre-analytical procedures, including information on sample type, primary containers, centrifugation conditions, and long-term storage protocols. Developed by the International Society for Biological and Environmental Repositories (ISBER), its primary goal is to ensure that each biobank sample is accompanied by a standardized set of metadata. In contrast, BRISQ offers broader guidance for reporting, including both *in vivo* and *in vitro* factors such as pre-acquisition procedures, transport parameters, and quality assurance steps relevant to downstream analysis. An analysis of BRISQ’s implementation in high-impact journals revealed that pre-analytical factors were often poorly reported, with little improvement observed following the publication of the guidelines in 2011.^20^ These findings suggest that the BRISQ guidelines have not yet been widely adopted and may be overly ambitious as an initial step towards improving pre-analytical reporting in the scientific literature.

In this study, we aim to assess the quality of pre-analytical reporting in recent clinical biomarker research using biobank plasma and serum samples, focusing on studies from beginning of 2018 to February 2023. We developed a scoring system based on SPREC, supplemented by BRISQ elements, to evaluate the comprehensiveness of reporting. Our ultimate objective is to raise awareness of the importance of the pre-analytical phase and encourage the adoption of standardized reporting practices to mitigate biases that may lead to misleading results and conclusions in biomedical literature.

## Methods

### Article Selection

We included studies that at time of the searches end of February 2023 were officially published or added to pubmed between 1 January 2018 and 28 February 2023 and utilized biobank plasma or serum samples for biomarker analysis. Articles were initially selected through a combination of search terms in the PubMed database, ensuring that the selected papers met the criteria of being published in English and focusing on plasma, serum, or urine samples, as outlined in Supplemental Table I. Studies were excluded if they met any of the following criteria: a) focused on the design of future biobanks, b) provided general information about biobanks without reporting pre-analytical details, c) examined the effect of pre-analytical factors on measurement outcomes, d) were review articles, e) involved animal research, f) did not use plasma or serum samples, g) lacked full-text availability, h) were not published in English, or i) did not explicitly mention biobanking or sample storage. Studies published before 2018 were also excluded, and for feasibility reasons of our literature study, we excluded studies involving urine as a sample type.

### Data Extraction

The selected articles were evaluated based on their reporting of various pre-analytical factors. These factors were derived from the SPREC guidelines and supplemented with additional elements from BRISQ. We included all relevant SPREC elements pertaining to plasma and serum as these focused on preanalytical processes for biobank biomaterials, and extended the evaluation by incorporating BRISQ elements such as clinical characteristics, storage duration, transport conditions, fasting status, separation gel use, and freeze-thaw cycles.^16-18^ To ensure objectivity, the data extraction process was piloted by the authors to minimize inter-observer bias.

Pre-analytical elements from SPREC and BRISQ were identified in the main article, supplementary materials, and references. The elements were categorized and recorded according to their presence and source of reporting. The final list of elements included internal transport, biospecimen type, fasting duration, demographic characteristics (age, sex), blood tube additives, separation gel, delays prior to and after centrifugation, centrifugation parameters (speed, time, temperature), storage conditions, and freeze-thaw cycles. Demographic details were recorded separately from the pre-analytical process. For each article, additional descriptive information was collected, including PubMed article ID, publication year, country of affiliation of the primary authors, journal name, impact factor (obtained from Clarivate), biobank name, use of multiple pre-analytical protocols, presence of external transport, and analysis purpose (Table 1).

**Table 1.** Characteristics and pre-analytical variables analysed in selected articles and their description. For the elements the source of the description was reported (main article, first line reference, or supplement)

| Variable | Description |
| --- | --- |
| **Descriptive Characteristics** | **Description of characteristics** |
| Article ID | PMID of the article, for identification |
| Year of publication | Year of publication in print |
| Country | Country of affiliation of the majority of authors, if there is a draw, the last author weighs heaviest |
| Journal | Journal in which the article was published. |
| Impact factor | Impact factor of the journal, found at [Journal Citation Reports - Home (clarivate.com)](https://www.clarivate.com). From the year the publication was published or the most recent one before if that year is not yet available |
| Biobank name | Name(s) of the biobank(s) used |
| Protocol | Use of multiple protocols during the study (yes or no) |
| External transport | Presence of external transport of the samples to different locations (yes or no) |
| Analysis purpose | Biomarker type that was measured in the samples |
| Biospecimen type | Presence of description of the biospecimen type (yes or no), and if so, if it was plasma or serum or both |
| **Pre-analytical elements** | **Description of elements** |
| Internal transport | Mention of temperature, or means of transport |
| Biological variables | Mention of fasting or non-fasting |
| Demographic characteristics | Mention of sex and age of the participants |
| Additive | Mention of the anticoagulant, for instance EDTA, heparin or citrate, or no anticoagulant (serum). |
| Separation gel | Mention of separation gel or no gel in the primary tube. If information about the type of tube is given but not the gel, and it is public knowledge that this type of tube always contains a gel, or never contains a gel this counts |
| Pre centrifugal delay | Mention of the delay between the collection and the moment of centrifuging is reported |
| Post centrifugal delay | Mention of the delay between the centrifuge step and the long-term storage. Total delay between collection and storage counts |
| Centrifuge time | Mention of duration of centrifuging |
| Centrifuge speed | Mention of speed of centrifuge |
| Centrifuge temperature | Mention of temperature during centrifuging |
| Long-term storage temperature | Mention of temperature of long-term storage |
| Inclusion period | Mention of time range in which the samples were collected, processed and stored |
| Analysing period | Mention of time period during which the samples are taken out of storage and analysed |
| Freeze-thaw cycle | Mention of the number of freeze thaw cycles that the samples went through |

### Scoring and Quality Control

Data extraction was performed by a single investigator (J.J.), with findings discussed in meetings involving all four authors: a master’s student in Medical Biology (J.J.), a professor of clinical chemistry (D.S.), an associate professor in epidemiology (F.V.), and the sample process coordinator from the Radboud Biobank (M.B.). Scoring was initially completed on paper and later transferred into Excel. Quality assessments were conducted in two rounds to minimize subjectivity across the dataset. After the initial review by the main assessor (J.J.) of 100 articles, 15 randomly selected papers were reviewed by two different other authors.

Each of the other authors (D.S, F.V., M.B) re-examined 10 articles. For the second round, 9 additional papers from the remaining 194 articles were reviewed by two other authors, with each of three other author re-examining 6 papers.

### Statistical Analysis

Descriptive statistics were performed using SPSS version 28 in all articles and in subgroups (papers on UK biobank, and papers with specific analysis purposes: miRNA and transcriptomics, metabolites, lipid metabolites and metabolomics measurements). We calculated the number of 14 specific pre-analytical elements reported for each article and presented the results as percentages across different continents, publication years, and biobanks. We also examined correlations between the number of reported elements, journal impact factor, and publication year using Spearman’s correlation coefficient. Inter-rater reliability was assessed using Fleiss’ kappa, and discrepancies in scoring were discussed, after which each individual author could adapt their scoring.

## Results

A total of 756 records were identified through the PubMed search. After applying the inclusion and exclusion criteria, 307 articles remained for data extraction. Following a further round of exclusions during data extraction, 294 articles were included in the final analysis (Figure 1). The 294 articles originated from 30 countries, were published in 194 journals, and involved 153 biobanks. The most frequently used biobank was the UK Biobank, which accounted for 23 (7.8%) of the articles (Table 2). The majority of studies were based in Western nations, with Europe (n=174, 59.2%), the Netherlands (n=33, 11.2%) and the United States (14.3%) being the largest contributors (Supplemental Table II). Among the 294 articles, 55.4% (n=163) focused on protein biomarkers, 20.4% (n=60) on metabolites, and 9.5% (n=28) on miRNA, with the remainder studying transcriptomics (n=5, 1.7%) or other biomarker types (Supplemental Table III). Detailed information on article characteristics is provided in Table 2 and Supplemental Tables II and III.

**Table 2.**
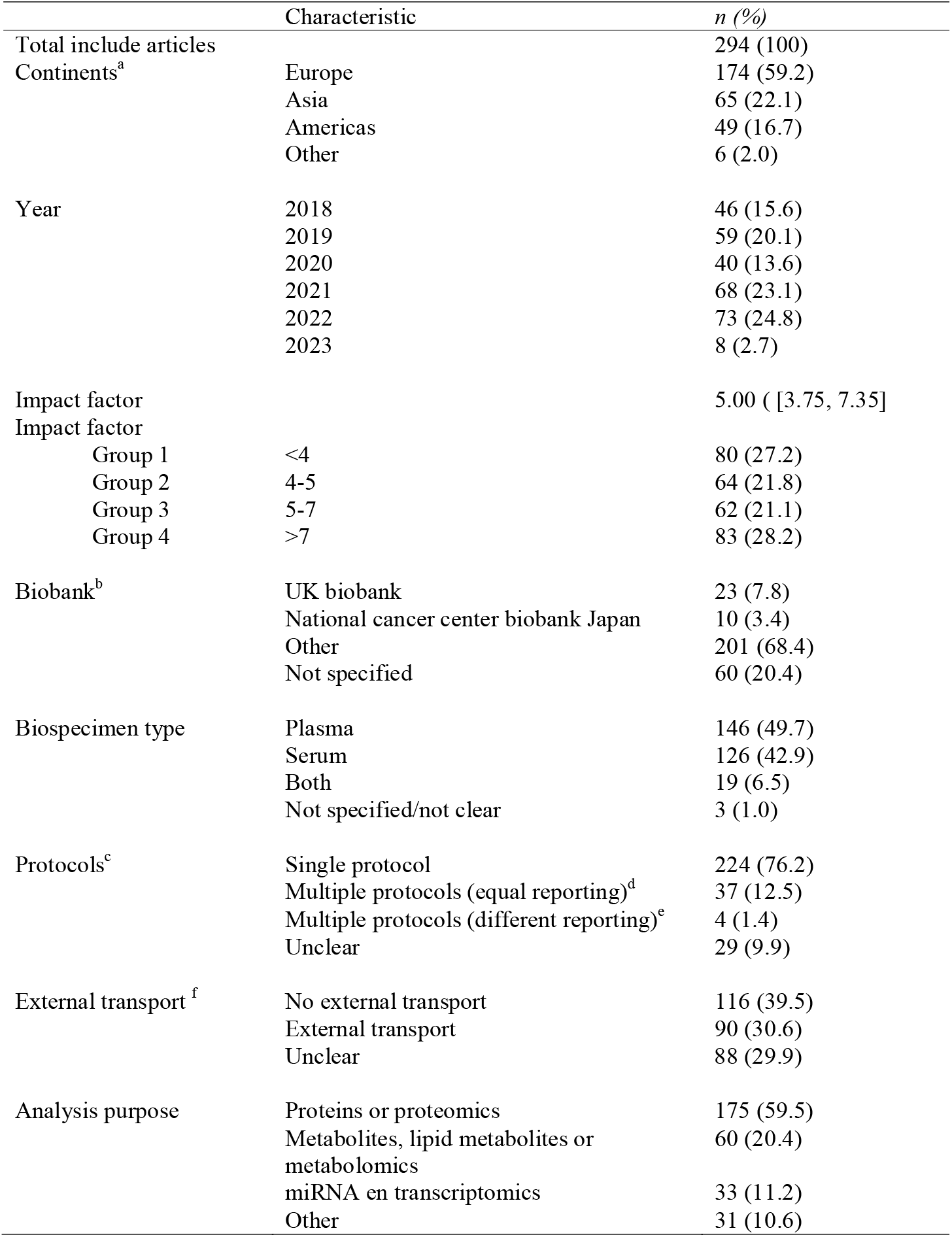
Characteristics of included articles that performed clinical biomarker research on biobank plasma or serum samples. Data are presented as n (%) or median [25^th^, 75^th^ percentile]. ^a^Continents of origin of the authors (List of specific countries can be found in supplement Table III). ^b^ Biobanks used in more than 6 papers; Others, biobank used in 6 papers or less. ^c^ One or multiple pre-analysis protocols described in the article.d same pre-analytical elements reported for the different protocols; ^e^, different pre-analytical elements reported for the different protocols; f Was there external transport from collection location to either a processing or storage location.

**Figure 1.**
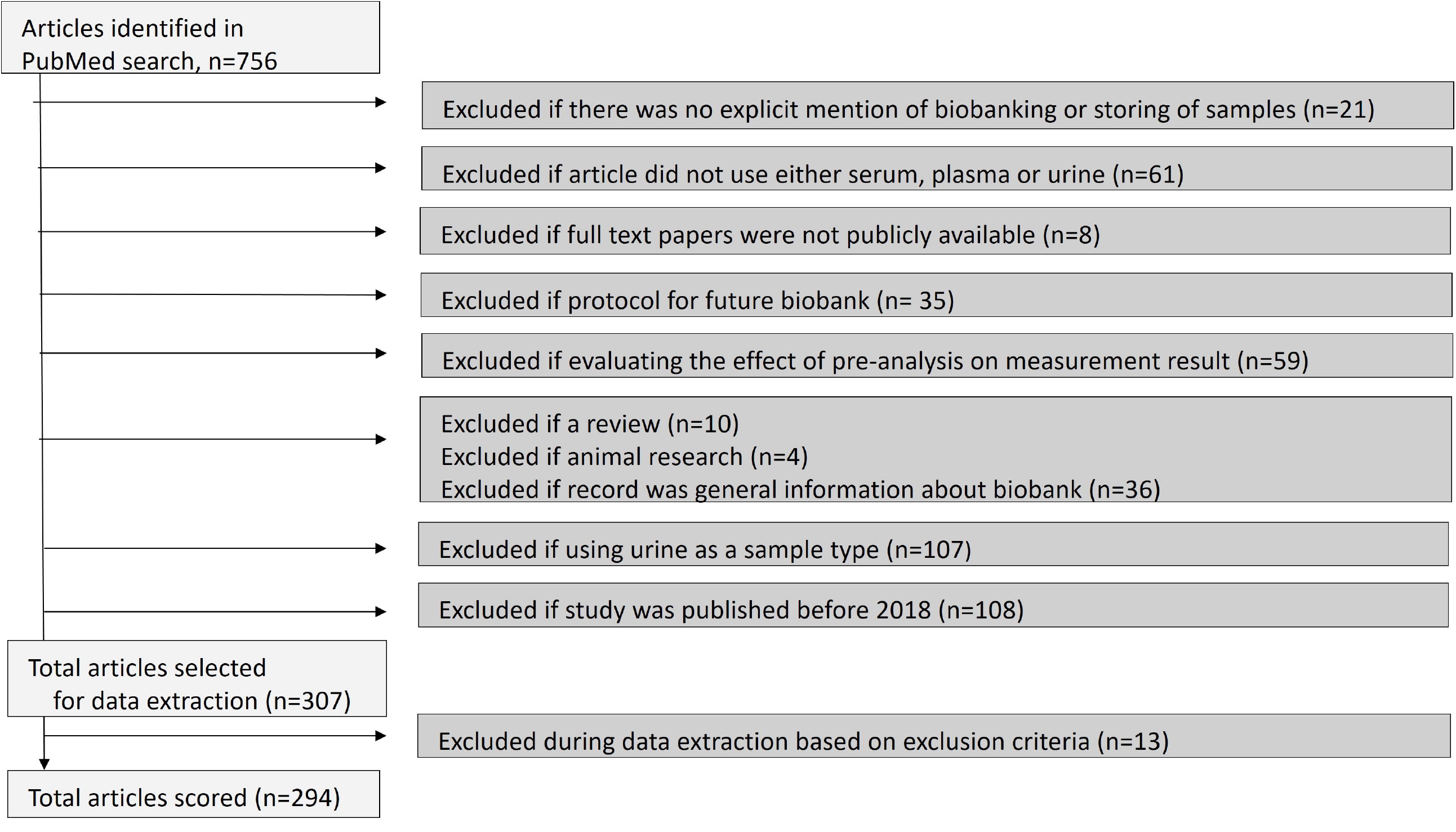
Flow diagram describing selection of articles. End of February 2023 we searched PUBMED for articles that made use of biobank serum, plasma or urine samples, and published between January 2018 and February 2023. Exact search terms that resulted in the 756 articles can be found in supplemental Table I. During initial selection we also exclude studies that used urine for biomarker studies and studies that were published before 2018. Subsequent use of selection criteria before and during data extraction resulted in 294 articles included for scoring in the study.

### Reporting of Pre-Analytical Elements

The average reporting of the pre-analytical elements listed in Table 1 across the 294 articles reviewed was 38.8% (Table 3). The reporting varied, with some elements being reported more frequently than others (Figure 2). Demographic characteristics, gender and age, were reported most consistently, i.e. in 96.9% of articles. Blood tube additives, long-term storage time, and the inclusion period were documented in 82.7%, 81.0%, and 72.4% of the articles, respectively. In contrast, all other pre-analytical elements were reported in fewer than 40% of articles. Internal transport, blood tube separation gel, pre- and post-centrifugal delays, freeze-thaw cycles, and the analysis period were mentioned in only 8.5%, 11.9%, 22.4%, 17.0%, 22.8%, and 8.8% of articles, respectively. The reporting of centrifugal conditions ranged between 20-35%, with centrifugal temperature, time, and speed reported in 20.7%, 33.0%, and 34.4% of articles, respectively. Fasting time was described in 31.0% of the articles (Figure 2, Table 3).

**Table 3.** Percentage of pre-analytical elements reported for all articles and subgroups. Data presented as percentage of articles mentioning element in either main article, supplement, or reference. *, for the UK biobank we found a relatively high between paper variation in referring to other reports for the pre-analytical elements as well as in the pre-analytical information provided in these articles (data not shown).

| Elements | All articles (%) | Articles using UK biobank* (%) | Articles measuring miRNA and transcriptomics (%) | Articles measuring metabolomics metabolites, and lipid metabolites (%) | Articles measuring proteins or proteomics (%) |
| --- | --- | --- | --- | --- | --- |
| Total (number, %) | 294 (100) | 23 (7.8) | 33 (11.2) | 60 (20.4) | 175 (59.5) |
| Internal transport | 8.5 | 30.4 | 0.0 | 13.3 | 7.4 |
| Fasting time | 31.0 | 47.8 | 6.0 | 45.0 | 29.7 |
| Demographics | 96.9 | 95.7 | 96.9 | 96.7 | 97.1 |
| Additives | 82.7 | 95.7 | 93.9 | 83.3 | 80.6 |
| Separation gel | 11.9 | 34.8 | 9.1 | 16.7 | 10.8 |
| Pre-centrifugal delay | 22.4 | 26.1 | 9.1 | 16.7 | 25.1 |
| Post-centrifugal delay | 17.0 | 34.8 | 15.2 | 15.0 | 18.9 |
| Centrifuge time | 33.0 | 30.4 | 27.2 | 23.3 | 35.9 |
| Centrifuge speed | 34.4 | 30.4 | 27.2 | 23.3 | 37.7 |
| Centrifuge temperature | 20.7 | 34.8 | 18.2 | 20.0 | 20.6 |
| Long term storage temperature | 81.0 | 52.2 | 81.8 | 70.0 | 85.1 |
| Inclusion period | 72.4 | 91.3 | 60.6 | 85.0 | 69.7 |
| Analyzing period | 8.8 | 17.4 | 3.0 | 5.0 | 12.0 |
| Freeze-thaw cycle | 22.8 | 30.4 | 24.2 | 31.7 | 18.3 |
| Average of elements | 38.8 | 46.6 | 33.8 | 38.9 | 38.7 |

**Figure 2.**
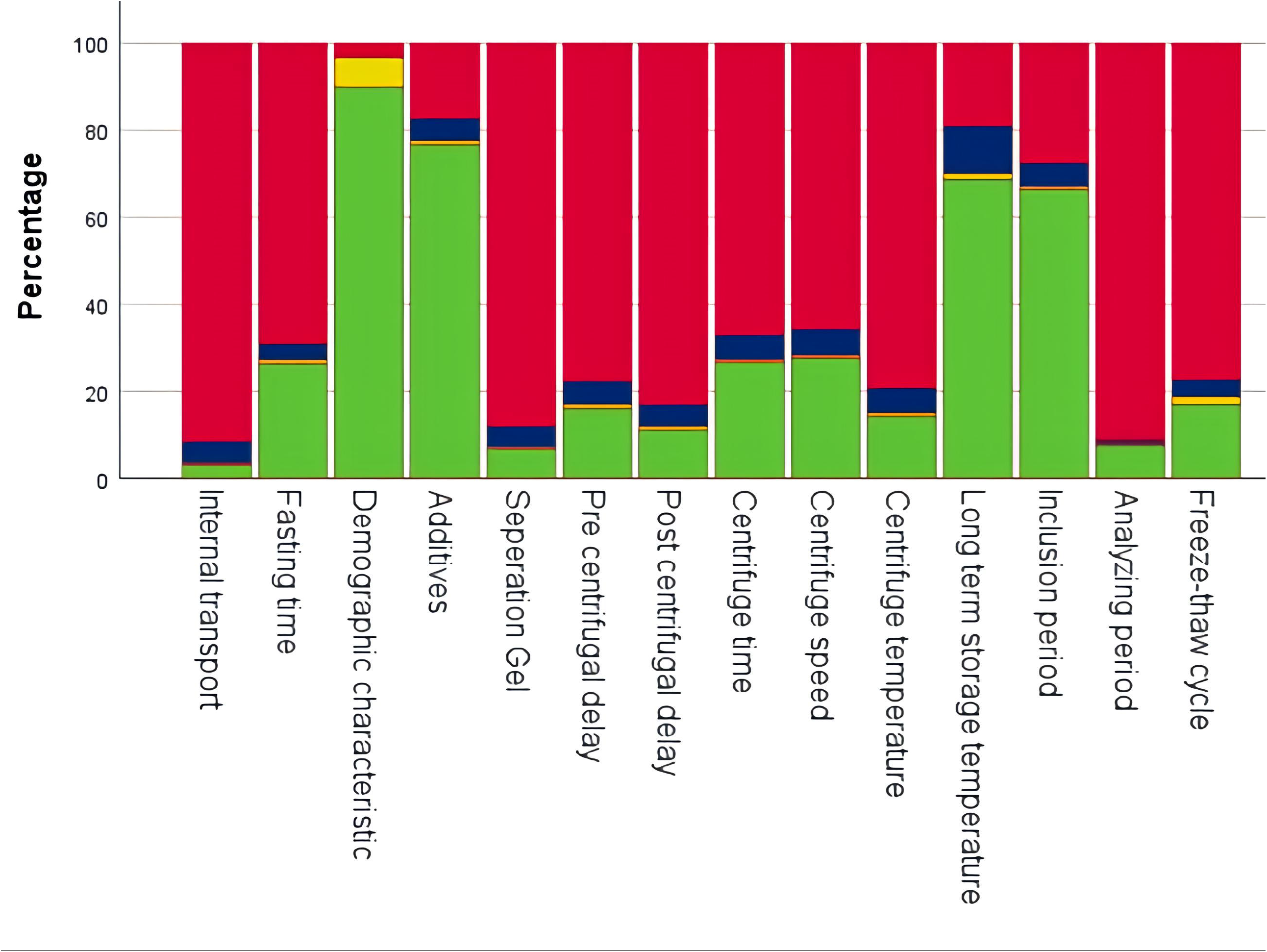
Reporting of pre-analytical elements. Reporting is expressed as percentage of the selected 294 articles that not reported (red) or reported a pre-analytical elements in main article text (green), supplement (yellow) or first line references (blue)

Among the reported elements, most were described in the main article (84.2%), while a smaller proportion appeared in the first-line references (12.6%) or the supplement (3.2%). The percentage of reporting in the main article varied by element, with additives, demographics, long-term storage temperature, and inclusion period reported in 76.5%, 89.8%, 68.7%, and 66.3%, respectively (Figure 2). To assess whether the percentage of reported pre-analytical elements differed for frequently used biobanks, we analyzed data from the UK Biobank, the most used biobank in our study, and compared these with the overall group. Of the 23 articles utilizing the UK Biobank, the average number of reported elements (46.6%) was higher than that of the entire group (38.8%, Table 3). However, the pre-analytical elements for the UK Biobank were more often cited in references than in the main articles, with centrifugal elements described in a reference in 27.5% of publications, compared to only 4.3% in the main articles. For all other articles, centrifugal elements were reported in 5.9% of the main articles and 22.8% in references (data not shown). Furthermore, there was considerable variability between UK-biobank articles in the pre-analytical information provided in references, with in some articles references containing comprehensive pre-analytical details, while references in other articles provided little to no relevant information (data not shown).

We found no correlation between the impact factor of journals and the total number of pre-analytical elements reported per article (Spearman’s correlation coefficient: 0.04). Similarly, there was no significant association between the year of publication and the number of elements reported (Spearman’s correlation coefficient: 0.05) (Supplemental Figures IA, 1B, and Table IV).

Given that miRNA, transcriptomics, and metabolites are particularly vulnerable to pre-analytical handling variations, we conducted a separate analysis of these analysis types. Despite the vulnerability of these analyses, pre- and post-centrifugal delays were reported less frequently or similarly for miRNA and transcriptomics (9.1% and 15.2%, respectively), and for metabolites (16.7% and 15.0%), compared to proteins/proteomics (25.1% and 18.9%) and the entire group of articles (22.4% and 17.0%, Table 3). Additionally, fasting time, which has been shown to impact metabolite measurements, was reported in 45% of metabolite articles, a higher percentage compared to articles on proteins (29.4%) and all articles (31.0%). Nevertheless, this still represents a minority of articles.

### Quality Assessment

The first quality assessment, conducted after the data collection of the first 100 articles, showed substantial to almost perfect agreement between each evaluator and the main assessor (Table 4). Following discussions of each article, overall agreement improved, with Fleiss’ Kappa measures ranging between 0.90 and 0.98. The second quality assessment, conducted after reviewing the subsequent 194 articles, demonstrated almost perfect agreement between evaluators and the main assessor. Fleiss’ Kappa values were 0.85, 0.83, and 0.80 for the main assessor (J.J.) and evaluators M.B., F.V., and D.S., respectively, with post-discussion values of 0.95, 0.88, and 0.94 (Table 4).

**Table 4.**
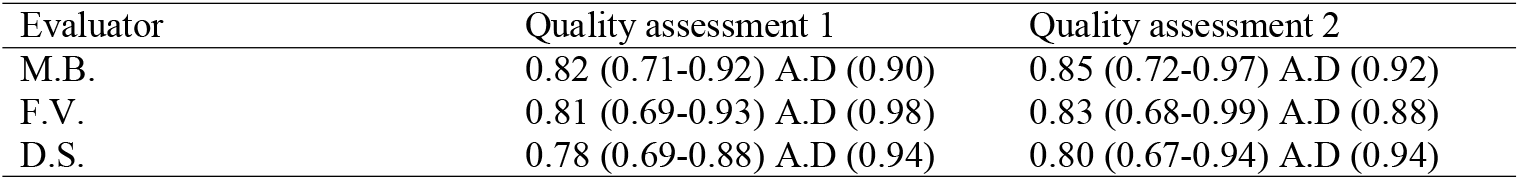
Quality assessment of agreement between evaluators. Agreement data are presented as Fleiss Kappa measure (95% confidence interval) between the main assessor (J.J.) and the three evaluators (M.B., F.V and D.S.) for 2 assessment rounds. Quality assessment 1 was performed after the first scoring round of 100 articles, and assessment 2 after scoring of the final 194 articles. A.D, after discussion: the Fleiss Kappa score after each evaluator was given the option to adapt the scores after discussing the discrepancies.

## Discussion

This study investigated the reporting of pre-analytical procedures for biobank samples used in biomarker research, based on a selection of SPREC and BRISQ criteria, in articles published between beginning of 2018 and end of February 2023. Our findings indicate that the reporting of pre-analytical elements is insufficient in the majority of articles, with many elements not reported or only partially described.

Demographic characteristics, blood tube additives, long-term storage time, and the inclusion period were relatively well documented, while other elements, such as fasting time, centrifugal conditions, freeze-thaw cycles, transport, and pre- and post-centrifugal delays, were poorly reported. The differences in reporting between different types of analysis were minimal, and no significant correlation was found between impact factor and the number of reported pre-analytical elements.

Our assessment focused on pre-analytical reporting in biobanking, using SPREC as a foundation for the selection of elements, supplemented with relevant BRISQ criteria.^16,18,19^ We believe that this framework provided an appropriate list of elements for reporting pre-analytical processes in biobanks. The findings of poor pre-analytical reporting align with those of a previous evaluation conducted a decade ago, which highlighted the limited implementation of BRISQ guidelines and the lack of improvement in reporting biospecimen data in scientific literature since their publication.^20^ The authors of this evaluation attribute this low uptake of BRISQ in scientific literature to its broadness, openness for interpretation, and the vague definition of some categories.

Our results demonstrate that, even today, most pre-analytical processes are reported in fewer than 25% of articles, which contrasts sharply with the near-universal reporting of demographic characteristics. This suggests a clear consensus in clinical research on the importance of documenting subject characteristics^21^, yet no similar consensus exists for pre-analytical processes. It is important to note that a lack of information on certain pre-analytical aspects does not necessarily imply that the pre-analysis is unsuitable for its intended purpose, but it does limit the ability to assess the reliability and robustness of the study’s conclusions.

We observed that the reporting of pre-analytical elements was not influenced by the type of analysis. Also articles on metabolites, miRNA and transcriptomics analyses demonstrated poor reporting of pre-centrifugal delays, despite evidence that delays in sample processing can alter the composition of the sample, potentially affecting these analysis outcomes.^12,22-24^ This is particularly the case when blood cells have not been separated from plasma or serum. Similarly, the reporting of fasting time was found to be suboptimal even for articles on metabolomic analysis, despite its known impact on these measurements. Our analysis found no relationship between the impact factor of journals and the completeness of pre-analytical reporting, which is concerning given that higher-impact journals are generally regarded as more influential on clinical decisions and guidelines. This suggests that the quality of reporting on pre-analytical procedures is not necessarily related to journal prestige, a factor that could undermine the credibility of research published in such journals.

Our study also highlighted differences in the reporting of pre-analytical elements for articles using the UK Biobank. Compared to the broader set of articles, for information on a higher percentage of pre-analytical elements UK biobank articles referred to the references rather than providing this information in the main text or supplementary materials. Interestingly, the majority of these references contained limited or no useful information about pre-analytical processes, with a minority that comprised all relevant pre-analytical information. These findings suggest that even when relevant information is available for a biobank, researchers may not be aware of its existence or importance.

We argue that complete reporting of pre-analytical procedures is a shared responsibility among the research community, including researchers, biobank organizations, funders and journal editors. We recommend that authors ensure the adequate reporting of pre-analytical processes, and that biobanks make this information readily accessible, for example through publicly available and regularly maintained websites.

The strength of our study lies in the large number of recent articles included, which were selected through a thorough search strategy. This enabled us to capture a broad picture of pre-analytical reporting in the biomedical field. Our study also comprises several limitations. First, as depicted in the flow diagram describing selection of articles, we slightly changed our study aim after the initial searches in which we also included urine as a biomaterial. Since this resulted in a number of articles, that exceeded our scorings capacity, at second instant, we decided to exclude articles that had urine as a sample type (rather than shortening the inclusion period). Second, by not using the search term ‘biorepository’ as a alternative word for ‘biobank’ we may have missed articles. Third, our searches did not select articles on analyte or ‘multi-analyte’ (omics) measurements in biobank materials that did NOT mention the term ‘biomarker’. Using this research strategy we may have missed articles on certain analytes measured in biobank biomaterials, among which are studies on cytokines. For reasons of feasibility of our literature search we also excluded GWAS studies, while these studies also use measurement results for certain analytes measured in biobank plasma or serum. We believe the abovementioned 2^nd^ and 3rd limitations have not affected our overall conclusion of poor reporting of pre-analysis in biomarker studies using biobank materials. However, they preclude to obtain some specific insights, for instance on reporting of pre-analysis in GWAS studies and of analytes typically not described by the term ‘biomarker’. Fourth, all analyses were conducted by a single assessor, although two rounds of quality assessments were carried out to mitigate this interobserver bias.

We envisage the list of pre-analytical elements that should be minimally reported in publications is a trade-off between transparency on how pre-analysis was conducted for elements that makes the sample fit for its intended use and the bureaucratic burden for researchers, funding agencies and article reviewers. It is beyond the scope of this study to propose a set of pre-analytical characteristics that meets these requirements, but we suggest it consists of a simple checklist approach for elements as outlined by SPREC, and BRISQ and that are adapted for differences in intended use. Better biospecimen data reporting increases transparency that is essential to evaluate credibility of scientific claims. Reporting may be facilitated by making pre-analytical biospecimen data available to the researcher at time of sample issuance. To this end, we encourage biobanks and suppliers to collaborate on inclusion of pre-analytical sample characteristics as described in SPREC and BRISQ standards in their Laboratory Information Management Systems (LIMS) and/or Biobank Information Management Systems (BIMS).

Improving quality of scientific publications by promoting transparent and accurate reporting is also the mission of the international network for Enhancing the QUAlity and Transparency Of health Research (EQUATOR). Its website showcases reporting guidelines for many study types, among which are CONSORT for randomized trials, and STARD for diagnostic and prognostic studies. Its library also comprises the BRISQ guideline for ‘biospecimen reporting’. However, to the best of our knowledge none of the biomedical journals requires adherence to the BRISQ (or SPREC) guideline for reporting pre-analytical variables for biomarker studies. We propose that journals that publish analyte measurement results of biobank samples refer to guidelines that provide guidance on reporting of the pre-analytical characteristics of biobank samples for clinical biomarker research.

Our findings underscore the importance of improving pre-analytical reporting in clinical biomarker research, as inadequate reporting can lead to irreproducible results and faulty conclusions, contributing to research waste. We call on researchers, biobank organizations, academic institutions, policymakers, funders and journal editors to prioritize transparent and accurate reporting of pre-analytical processes and to collaboratively develop and implement reporting guidelines that address the specific needs of different analysis purposes.

## Data Availability

All data produced in the present study are available upon reasonable request to the authors

## Authorship Contribution

D.S. conceived the study. D.S., F.V., and M.B. designed the study. All authors contributed critical insights into the search strategy, article selection, data extraction, interpretation, and analysis. J.J. performed the search and data selection. D.S., F.V, and M.B. screened a selection of articles for quality assessment. J.J. wrote the first draft of the manuscript. D.S. wrote subsequent versions of the manuscript with J.J., F.D., and M.B. providing essential input for revisions. All authors had full access to all data and were responsible for the decision to submit the manuscript for publication.

## Research Funding

Non declared

