## Supplemental material for "Reporting of pre-analytical Processes in Biobanked Biomaterials: A Missed Opportunity to Address the Replicability Crisis in Clinical Biomarker Research"

**Supplemental Table I. Search filters used for article collection in PubMed**

| | **Full search PubMed** | **Description** | Results |
|---|---|---|---|
| 1 | "Biomarkers"[Mesh] OR biomarker*[tiab] | Biomarkers | 1,092,882 |
| 2 | "Biological Specimen Banks"[Mesh] OR biobank*[all fields] OR bio-bank*[tiab] | Biobank | 34,154 |
| 3 | ("Biomarkers"[Mesh] OR biomarker*[tiab] ) AND ("Biological Specimen Banks"[Mesh] OR biobank*[all fields] OR bio-bank*[tiab]) | 1 AND 2 | 3,383 |
| 4 | ("Biomarkers"[Mesh] OR biomarker*[tiab] ) AND ("Biological Specimen Banks"[Mesh] OR biobank*[all fields] OR bio-bank*[tiab]) AND ("Plasma"[Mesh] OR plasma[tiab] OR "urine"[MeSH Subheading] OR "urine"[tiab] OR "urine"[MeSH Terms] OR "Serum"[Mesh] OR serum[tiab]) | 3 AND serum or plasma or urine | 979 |
| 5 | ("Biomarkers"[Mesh] OR biomarker*[tiab] ) AND ("Biological Specimen Banks"[Mesh] OR biobank*[all fields] OR bio-bank*[tiab]) AND ("Plasma"[Mesh] OR plasma[tiab] OR "urine"[MeSH Subheading] OR "urine"[tiab] OR "urine"[MeSH Terms] OR "Serum"[Mesh] OR serum[tiab]) NOT (Animals[MeSH] NOT Humans[MeSH]) | 4 AND human (not animal) | 965 |
| 6 | ("Biomarkers"[Mesh] OR biomarker*[tiab] ) AND ("Biological Specimen Banks"[Mesh] OR biobank*[all fields] OR bio-bank*[tiab]) AND ("Plasma"[Mesh] OR plasma[tiab] OR "urine"[MeSH Subheading] OR "urine"[tiab] OR "urine"[MeSH Terms] OR "Serum"[Mesh] OR serum[tiab]) NOT (Animals[MeSH] NOT Humans[MeSH]) AND (2013:2022[pdat]) | 5 AND filter 2013-2022 | 859 |
| 7 | (("Biomarkers"[Mesh] OR biomarker*[tiab] ) AND ("Biological Specimen Banks"[Mesh] OR biobank*[all fields] OR bio-bank*[tiab]) AND ("Plasma"[Mesh] OR plasma[tiab] OR "urine"[MeSH Subheading] OR "urine"[tiab] OR "urine"[MeSH Terms] OR "Serum"[Mesh] OR serum[tiab]) NOT (Animals[MeSH] NOT Humans[MeSH]) AND (2013:2022[pdat])) NOT (review[Publication Type]) | 6 NOT review | 821 |
| 8 | ((((("Biomarkers"[MeSH Terms] OR "biomarker*"[Title/Abstract]) AND ("Biological Specimen Banks"[MeSH Terms] OR "biobank*"[All Fields] OR "bio bank*"[Title/Abstract]) AND ("Plasma"[MeSH Terms] OR "Plasma"[Title/Abstract] OR "urine"[MeSH | 7 AND English(language) | 816 |

| | | | |
|---|---|---|---|
| | Subheading] OR "urine"[Title/Abstract] OR "urine"[MeSH Terms] OR "Serum"[MeSH Terms] OR "Serum"[Title/Abstract])) NOT ("animals"[MeSH Terms] NOT "humans"[MeSH Terms])) AND 2013/01/01:2022/12/31[Date - Publication]) NOT "review"[Publication Type]) AND "english"[Language] | | |
| 9 | (((((("Biomarkers"[MeSH Terms] OR "biomarker*"[Title/Abstract]) AND ("Biological Specimen Banks"[MeSH Terms] OR "biobank*"[All Fields] OR "bio bank*"[Title/Abstract]) AND ("Plasma"[MeSH Terms] OR "Plasma"[Title/Abstract] OR "urine"[MeSH Subheading] OR "urine"[Title/Abstract] OR "urine"[MeSH Terms] OR "Serum"[MeSH Terms] OR "Serum"[Title/Abstract])) NOT ("animals"[MeSH Terms] NOT "humans"[MeSH Terms])) AND 2013/01/01:2022/12/31[Date - Publication]) NOT "review"[Publication Type]) AND "english"[Language]) NOT ("genome wide association study"[MeSH Terms] OR ("genome wide"[All Fields] AND "association"[All Fields] AND "study"[All Fields]) OR "genome wide association study"[All Fields] OR "gwas"[All Fields]) | 8 NOT genome wide association study | 758 |
| 10 | ((((((("Biomarkers"[MeSH Terms] OR "biomarker*"[Title/Abstract]) AND ("Biological Specimen Banks"[MeSH Terms] OR "biobank*"[All Fields] OR "bio bank*"[Title/Abstract]) AND ("Plasma"[MeSH Terms] OR "Plasma"[Title/Abstract] OR "urine"[MeSH Subheading] OR "urine"[Title/Abstract] OR "urine"[MeSH Terms] OR "Serum"[MeSH Terms] OR "Serum"[Title/Abstract])) NOT ("animals"[MeSH Terms] NOT "humans"[MeSH Terms])) AND 2013/01/01:2022/12/31[Date - Publication]) NOT "review"[Publication Type]) AND "english"[Language]) NOT ("genome wide association study"[MeSH Terms] OR ("genome wide"[All Fields] AND "association"[All Fields] AND "study"[All Fields]) OR "genome wide association study"[All Fields] OR "gwas"[All Fields])) AND (fft[Filter]) | 9 AND full text available | 756 |

**Note that later in the study we excluded urine as a biomaterial (Figure 1).**

**Supplemental Table II. Country of affiliation of the majority of authors for each article.**

| | Number | Percentage |
|---|---|---|
| Canada | 7 | 2.4 |
| China | 27 | 9.2 |
| Denmark | 11 | 3.7 |
| Finland | 6 | 2.0 |
| France | 7 | 2.4 |
| Germany | 23 | 7.8 |
| Italy | 9 | 3.1 |
| Japan | 16 | 5.4 |
| Korea | 7 | 2.4 |
| Norway | 6 | 2.0 |
| Spain | 15 | 5.1 |
| Sweden | 26 | 8.8 |
| The Netherlands | 33 | 11.2 |
| United Kingdom | 22 | 7.5 |
| United States of America | 42 | 14.3 |
| Total | 294 | 100.0 |

Country is defined as country of affinity of the majority of authors, if a draw occurred the country of the last author was selected.

**Supplemental Table III. Number (%) of articles using specific analysis purposes.**

| | Number | Percent |
|---|---|---|
| Protein | 163 | 55.4 |
| miRNA | 28 | 9.5 |
| Metabolomics | 25 | 8.5 |
| Selected metabolite | 22 | 7.5 |
| Lipid metabolites | 13 | 4.4 |
| Proteomics | 12 | 4.1 |
| Cell free DNA | 9 | 3.1 |
| Vitamin | 6 | 2.0 |
| Transcriptomics | 5 | 1.7 |
| Other omics | 4 | 1.4 |
| Other | 4 | 1.4 |
| Electrolyte | 2 | .7 |
| Cardiac markers | 1 | .3 |
| Total | 294 | 100.0 |

**Supplemental table IV.** Number of articles and pre-analytical elements reported per publication year and impact factor.

| | **Publication year** | | | | | | **Impact factor** | | | |
|---|---|---|---|---|---|---|---|---|---|---|
| | **2018** | **2019** | **2020** | **2021** | **2022** | **2023** | **<4** | **4-5** | **5-7** | **>7** |
| **Number of articles** | 46 | 59 | 40 | 68 | 73 | 8 | 82 | 64 | 62 | 83 |
| **Reported elements (mean, SD)** | 7.00, 0.41 | 6.53, 0.35 | 6.33, 0.52 | 6.96, 0.41 | 6.99, 0.31 | 7.88, 0.85 | 6.70, 0.32 | 6.50, 0.39 | 7.16, 0.33 | 6.93, 0.34 |

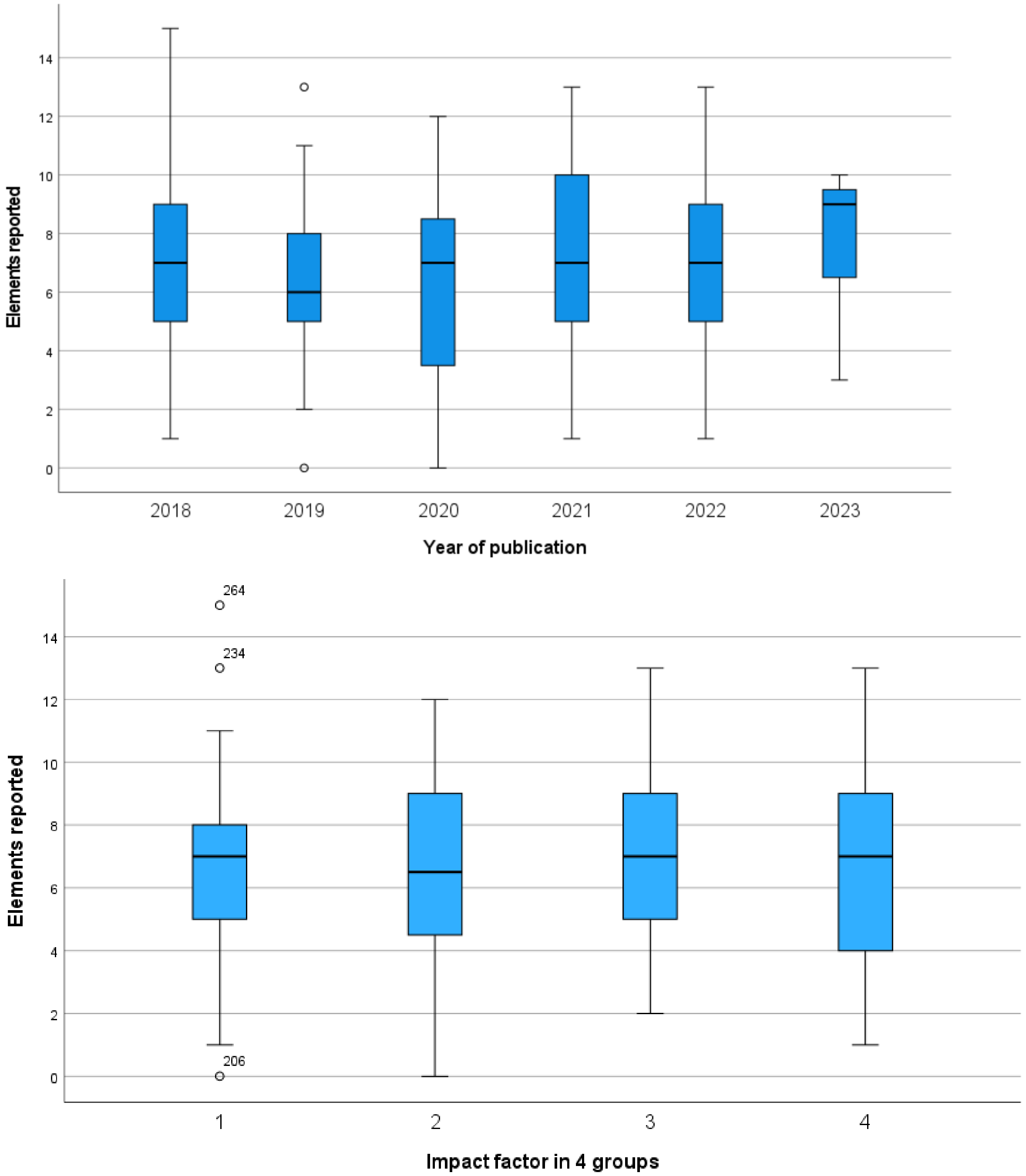


**Supplemental Figure I. Top:** Number of pre-analytical elements reported per publication year. Boxes indicate median and 25, 75% ranges; Whiskers describe the 95% confidence interval. Spearman correlation coefficient calculation showed a non significant correlation (Rho of 0.05).

**Bottom:** Number of pre-analytical elements reported for 4 groups of impact factors. Impact factor (IF) groups: 1= IF < 4; 2= IF 4-5; 3= IF 5-7; 4= IF> 7. Boxes indicate median and 25$^{th}$- 75$^{th}$ % ranges; Whiskers describe the 95% confidence interval. Spearman correlation coefficient calculation showed a non significant correlation (Rho of 0.05).
